# The comparability of Anti-Spike SARS-CoV-2 antibody tests is time-dependent: a prospective observational study

**DOI:** 10.1101/2021.08.26.21262426

**Authors:** Thomas Perkmann, Patrick Mucher, Nicole Perkmann-Nagele, Astrid Radakovics, Manuela Repl, Thomas Koller, Klaus G Schmetterer, Johannes W Bigenzahn, Florentina Leitner, Galateja Jordakieva, Oswald F Wagner, Christoph J Binder, Helmuth Haslacher

**Affiliations:** Department of Laboratory Medicine, Medical University of Vienna, Austria; Department of Physical Medicine, Rehabilitation and Occupational Medicine, Medical University of Vienna, Austria

**Keywords:** SARS-CoV-2, serology, vaccination, agreement, time-dependency

## Abstract

**Objectives:** Various commercial anti-Spike SARS-CoV-2 antibody tests are used for studies and in clinical settings after vaccination. An international standard for SARS-CoV-2 antibodies has been established to achieve comparability of such tests, allowing conversions to BAU/ml. This study aimed to investigate the comparability of antibody tests regarding the timing of blood collection after vaccination.

**Methods:** For this prospective observational study, antibody levels of 50 participants with homologous AZD1222 vaccination were evaluated at 3 and 11 weeks after the first dose and 3 weeks after the second dose using two commercial anti-Spike binding antibody assays (Roche and Abbott) and a surrogate neutralization assay.

**Results:** The correlation between Roche and Abbott changed significantly depending on the time point studied. Although 3 weeks after the first dose, Abbott provided values three times higher than Roche, 11 weeks after the first dose, the values for Roche were twice as high as for Abbott, and 3 weeks after the second dose even 5-6 times higher.

**Conclusions:** The comparability of quantitative anti-Spike SARS-CoV-2 antibody tests is highly dependent on the timing of blood collection after vaccination. Therefore, standardization of the timing of blood collection might be necessary for the comparability of different quantitative SARS-COV-2 antibody assays.

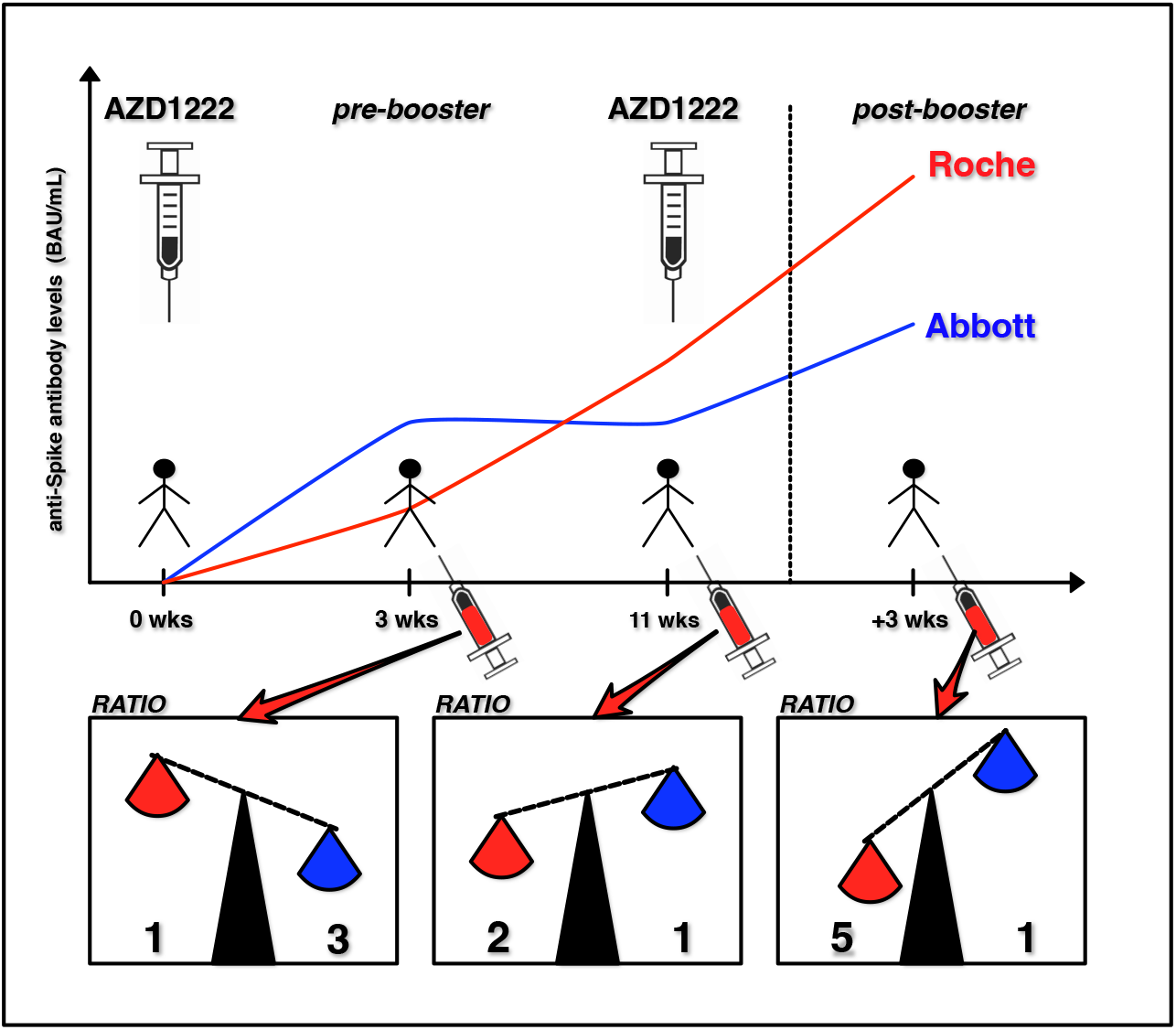

## Introduction

Infectious diseases continue to pose a significant challenge for humanity, as the SARS-CoV-2 pandemic has again demonstrated^1^. Nevertheless, in contrast to the past, diagnostic, therapeutic, and preventive strategies are now being developed at an unprecedented rate to address these pandemic challenges. Among all these strategies, however, one stands out: vaccination against SARS-CoV-2. Using new technologies and extensive knowledge on active immunization against numerous pathogens, highly efficient vaccines have been developed and applied within a few months^2^.

The vaccination aims to induce a SARS-CoV-2 specific immune response in analogy to a passed infection, which should protect against disease or even better against infection. The methodically simplest way to objectify a particular immune reaction is to measure infection- or vaccine-induced specific antibodies^3,4^. Thus, antibody tests have been used for SARS-CoV-2 to confirm prior infection or detect unreported infections as part of seroprevalence surveys^5–7^. Various antigens have been applied in this regard, which fall into two classes: SARS-CoV-2 nucleocapsid-specific antibodies and antibodies directed against the spike protein^8^. The latter antibodies, which are formed against components of the virus surface spike protein, are induced by all COVID vaccines currently in use, making them an ideal surrogate for the immune response after vaccination^9^.

The need to establish quantitative assays to detect vaccine-induced antibodies was pointed out early on^10^. Furthermore, an international standard for SARS-CoV-2 antibodies (NIBSC 20/136) was issued by the WHO to improve the comparability of such assays^11^. Although there is currently no general recommendation to determine antibody levels in all individuals after SARS-CoV-2 vaccination, this is reasonable from a scientific perspective in the search for a correlate of protection and has been done in numerous studies^12–19^. Moreover, it is now known that suboptimal or even lack of response to vaccination can occur in specific groups like immunocompromised patients^20–26^. These potential non-responders might be identified in a first step by determining the antibody levels after vaccination. Unfortunately, there is little scientific evidence on the real-life comparability of different commercially available quantitative test systems^27–29^.

We could previously show that referencing the WHO SARS-CoV-2 antibody standard by reporting standardized binding antibody units (BAU/mL) is insufficient for different test systems to provide numerically comparable results^27^. We demonstrated this in a strictly standardized study setting concerning the time of blood collection and the vaccine used: three weeks after the first dose of BNT162b2. It was expected and has already been shown that antibody responses are dependent on the type of vaccine used^30,31^. Moreover, time kinetics of post-vaccination antibody levels have been described for different vaccines and various antibody assays^17,32–37^. However, whether and how these influencing factors affect the comparability of different quantitative SARS-CoV-2 antibody tests has not been systematically investigated. But answering these questions is fundamental to finding pragmatic ways to compare results from various SARS-CoV-2 antibody tests.

In the present work, we aimed to expand knowledge on the comparability of quantitative anti-spike SARS-CoV-2 assays using another commonly administered vaccine, AZD1222, combined with antibody measurements at multiple time points: three weeks after the first vaccine dose, 11 weeks after the first dose (immediately before the second dose), and finally three weeks after the second dose. Moreover, pre- and post-booster levels were compared to SARS-CoV-2 specific T-cellular interferon γ responses. This design allowed us, utilizing two of the most commonly used commercially available assays applied in post-COVID vaccination antibody studies, the Roche Elecsys SARS-CoV-2 S-ECLIA^38–41^and the Abbott Anti-SARS-CoV-2 IgG II^22,42,43^, to examine in detail the comparability of the assays concerning the timing of blood collection after vaccination.

## Methods

### Study design and participants

We included sera of 50 participants in this prospective observational performance evaluation study. Inclusion criteria were an age ≥18 years and willingness to donate blood in the course of the MedUni Wien Biobank’s healthy blood donor collection (Medical University of Vienna ethics committee vote No. 404/2012). Incomplete follow-up samples and seropositivity for anti-nucleocapsid antibodies due to infection with SARS-CoV-2 lead to exclusion. The study protocol was reviewed and approved by the Medical University of Vienna ethics committee (1066/2021) and conforms with the Declaration of Helsinki.

### Laboratory methods

Blood samples were taken 3 weeks and 11 weeks (“pre-booster”) after the first dose of AZD1222, as well as 3 weeks after dose 2. At pre- and post-booster time points, an additional amount of 4mL blood was drawn to estimate T-cellular immunity (see Fig. 1). Blood samples were processed and stored according to standard operating procedures by the MedUni Wien Biobank in an ISO 9001:2015 certified environment ^44^.

**Fig. 1:**
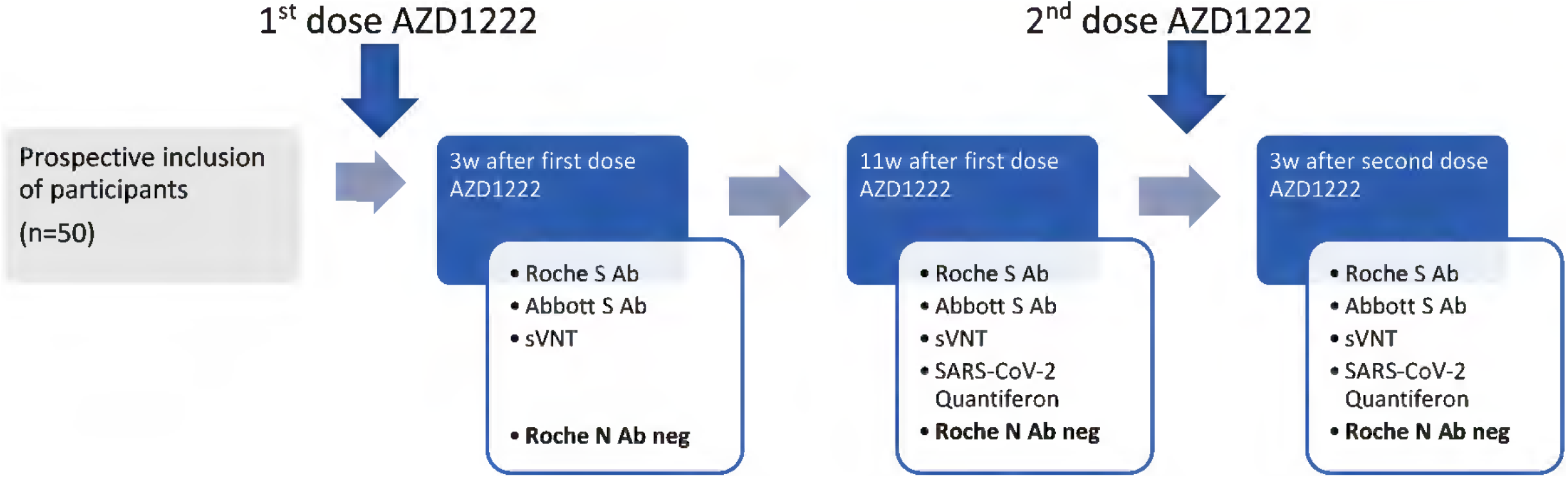
Study flow chart. Anti-Spike(S)-antibody (Ab) assays: Roche S, Abbott S. Infection with SARS-CoV-2 was ruled out by detection of antibodies against the SARS-CoV-2 nucleocapsid (N) using the Roche N ECLIA. W… weeks; sVNT… surrogate virus neutralization test.

Previous SARS-CoV-2 infection was assessed by the Roche Elecsys SARS-CoV-2 nucleocapsid ECLIA (Roche, Rotkreuz, Switzerland) on cobas e801 modular analyzers (Roche)^45^. This assay detects total antibodies against the viral nucleocapsid, which are induced after infection, but not after vaccination with AZD1222.

Vaccine-induced antibodies against the viral spike protein were quantified using the Roche Elecsys SARS-CoV-2 S-ECLIA^46^ and the Abbott Anti-SARS-CoV-2 IgG II^47,48^. This Roche test is a quantitative (range: 0.4 – 2,500 BAU/mL) total antibody sandwich assay recognizing antibodies directed against the receptor-binding domain (RBD) of the SARS-CoV-2 spike (S) protein and was performed on cobas e801 modular analyzers (Roche), samples >0.8 BAU/mL are considered diagnostically positive. As a deviation from the product manual, samples exceeding the quantification range were manually pre-diluted at a dilution factor of 1:10. The Abbott assay also quantifies anti-RBD specific IgG-antibodies (range: 1.0-11,360.0 BAU/mL) and was applied on an Abbott Architect i2000r (Abbott, USA). The assay’s threshold for positivity is 7,1 BAU/mL.

Binding reactivities (Roche, Abbott) were compared to a well-described CE-IVD marked surrogate virus neutralization test (GenScript cPass sVNT)^49–53^. This sVNT quantifies the serum’s ability to inhibit spike/ACE-2 interaction; results with an inhibition >30% are considered positive.

The T-cellular activity was estimated in pre- and post-booster whole blood samples using the Quantiferon SARS-CoV-2 assay (Qiagen, Hilden, Germany ^54–56^). In brief, we quantified the interferon (IFN)γ-release after 21-hour incubation of 1 mL heparinized whole blood portions with two different SARS-CoV-2 antigen mixtures (Ag1, Ag2), and with a negative (“Nil”) and a mitogen control, using an ELISA (Qiagen). For each patient and time point, IFNγ-values of the Nil control were subtracted from the SARS-CoV-2 specific antigen mixes results of the samples and presented as Ag1-Nil and Ag2-Nil, respectively.

### Statistical analysis

Continuous data are presented as median (interquartile range), and categorical data as counts (percentages). Paired data are compared by Wilcoxon- and Friedman-Tests. Correlations are calculated according to Spearman. Serological assays are compared by Passing-Bablok regressions, which assess differences between two test systems by estimating the slope (systematic proportional differences) and the intercept (systematic constant differences) of a linear regression line. There are no preconditions regarding the distribution of the measured values and the measurement errors to be met. P-values <0.05 were considered statistically significant. All calculations have been performed using MedCalc 19.7 (MedCalc bvba, Ostend, Belgium) and SPSS 26 (IBM, Armonk, USA). Graphs were drawn with Prism 9 (GraphPad, La Jolla, USA).

## Results

### Agreement between Roche and Abbott assays depends on the timing of blood collection

Blood samples of 50 individuals were collected 3 weeks and 11 weeks after the first dose of AZD1222 (11 weeks = “pre-booster”) and 3 weeks after the second dose (“post-booster”). Participant characteristics and binding assay levels at all assessed time points are presented in Tbl. 1 and Fig. 2.

**Tbl. 1:** Participant characteristics and surrogates of humoral (Roche, Abbott, sVNT) and cellular (IFNγ) immunity. P-values were derived from Friedman-tests. 3w… 3 weeks; BAU/mL… binding antibody units per milliliter; immunosuppr… immunosuppressive; sVNT… surrogate virus neutralization test; IFNγ… interferon γ

|  | Median (interquartile range) or counts (%) |  |  |  |
| --- | --- | --- | --- | --- |
| Female sex | 43 (86%) |  |  |  |
| Age [years] | 35.5 (29-49) |  |  |  |
| Immunosuppr. drugs | 2 (4%) |  |  |  |
|  | 3w post dose 1 | pre-booster | 3w post-booster | P-value |
| Roche [BAU/mL] | 13.6 (5.2-29.9) | 60.2 (36.4-112.8) | 895.5 (611.8-1681.0) | <0.0001 |
| Abbott [BAU/mL] | 42.2 (26.0-79.0) | 32.9 (20.8-53.7) | 171.2 (123.4-278.7) | <0.0001 |
| sVNT [% inhibition] | 47.1 (35.5-60.6) | 49.0 (35.3-62.4) | 95.2 (92.1-97.9) | <0.0001 |
| IFNγ Ag1-Nil [IU/mL] | | 0.10 (0.05-0.16) | 0.15 (0.09-0.32) | <0.0001 |
| IFNγ Ag2-Nil [IU/mL] | | 0.16 (0.09-0.31) | 0.31 (0.16-0.98) | <0.0001 |

**Fig. 2:**
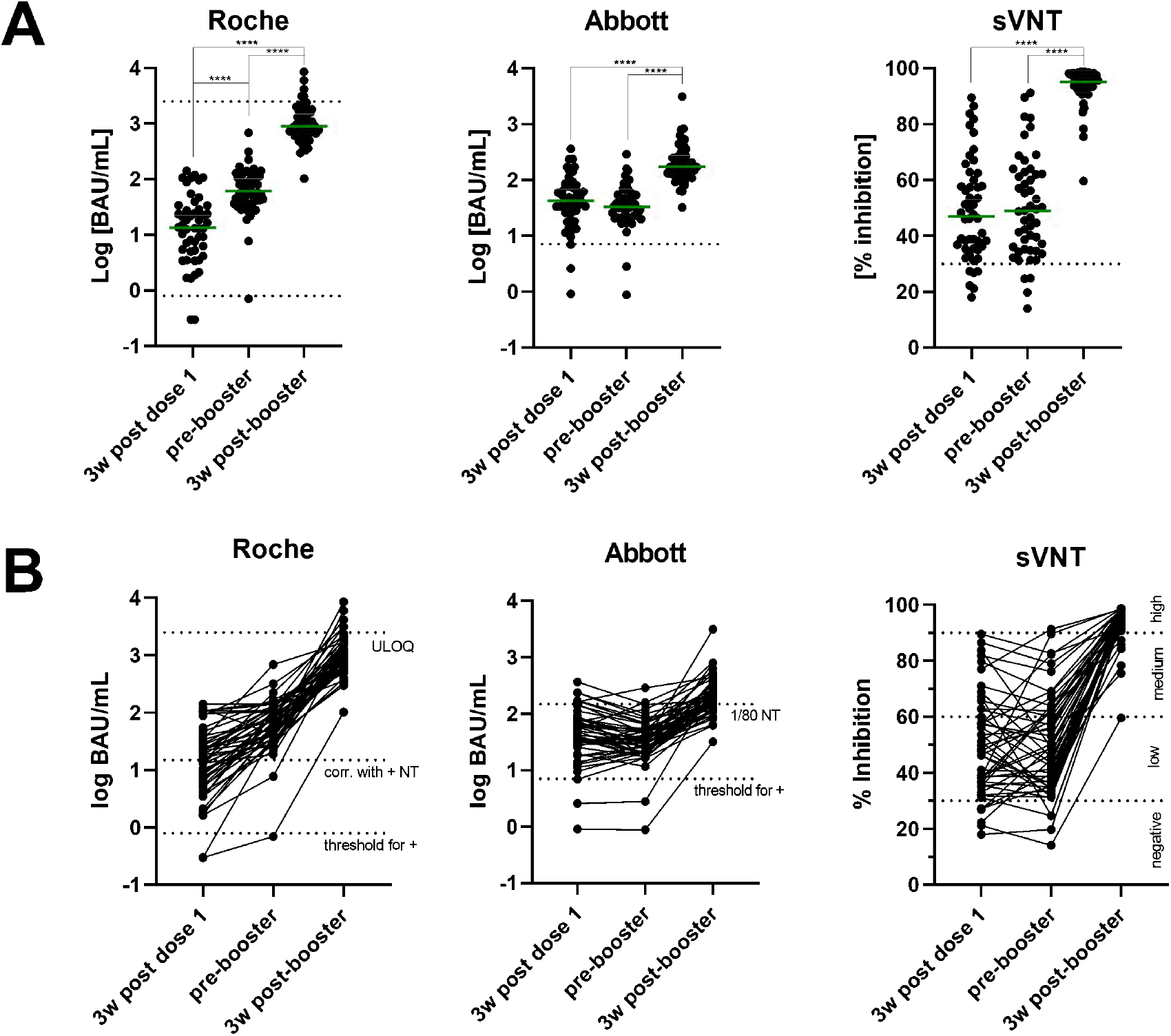
**A** Antibody levels (Roche, Abbott) and percent inhibition in a surrogate virus neutralization test (sVNT) 3w (weeks) after the first dose of AZD1222, pre-booster (11w after the first dose), and 3w post-booster. Dotted Lines indicate the test systems’ thresholds for positivity (Roche: 0.8 BAU/mL, Abbott 7.1 BAU/mL, sVNT 30%) and, in the case of Roche, the upper level of quantification (2,500 BAU/mL). Green lines represent the group median. ****… P<0.0001 in Wilcoxon tests. **B** Longitudinal changes of individual Roche, Abbott, and cPass surrogate virus neutralization test (sVNT) results: 3w (3 weeks) after the first dose, before the booster dose, and 3w after the booster dose. According to the manufacturers, Roche results ≥15 BAU/mL correlate with a positive neutralization test, Abbott results ≥149,1 BAU/mL correspond to a neutralization titer of at least 1:80; 30% inhibition is considered the sVNTs threshold for positivity. Results are, according to the manufacturer, categorized into low (30-60%), medium (60-90%), and high (>90%) neutralizing capacity (all levels indicated by dotted lines).

In brief, Roche S antibody levels significantly increased from 13.55 BAU/mL (5.21-29.88) at 3 weeks after the first dose to 60.20 (36.38-112.80) directly before the booster (all P<0.0001). 3 weeks after the booster, the median levels were 895.50 (611.80-1681.00). With Abbott, results remained stable between 3 and 11 weeks after the first dose: 42.23 BAU/mL (26.00-78.99) and 32.88 (20.78-53.69), P=0.178, and rose to 171.20 (123.40-278.70) 3 weeks after the booster (P<0.0001). Similar to the Abbott test, the sVNT did not show significant changes between week 3 and 11 after the first dose but significantly increased 3 weeks after the second dose: 47.1 % inhibition (35.5-60.6), 49.0 (35.2-62.4), 95.2 (92.1-97.9), P_3w post-booster vs. pre-booster or 3w after dose 1_<0.0001. In terms of relative changes in antibody levels for individual participants when comparing 3 and 11 weeks after the first dose versus 3 weeks after the second dose, we observed a 14.2 (8.4-30.8) and 80.8 [27.4-191.0] fold change in titers for Roche, and a 4.9 (3.0-10.1) and 4.5 [2.2-9.9] fold change for Abbott. Thus, the Roche test discriminated increases in antibody levels between weeks 3 and 11 after the first dose: 4.7 (2.2-9.5), whereas Abbott did not: 0.8 (0.5-1.4).

3 weeks after the first dose, results from Roche and Abbott binding assays showed a moderate correlation (ρ=0.755, P<0.0001). Passing-Bablok regression analysis revealed the following equation: Abbott = 7.4 + 2.99*Roche, whereby only the slope of the equation was statistically significant (2.06 – 17); intercept: −4.0 – 13.9). The agreement between both tests improved markedly 11 weeks after the first dose, with the correlation coefficient rising to ρ=0.902, P<0.0001. Passing-Bablok regression revealed that BAU/mL derived from Abbott were approximately half those measured by Roche: Abbott = 1.1 + 0.50 * Roche (intercept: −2.4 – 5.5, slope: 0.43 – 0.56). 3 weeks after dose 2, the correlation between Roche and Abbott results remained excellent (ρ=0.950, P<0.0001); however, the conversion between the values changed again, with Roche values approximately 5-6 times higher than Abbott results: Abbott = 13.3 + 0.178 * Roche (intercept: −4.5 – 29.5; slope: 0.16 – 0.20). (Figure 3)

**Fig. 3:**
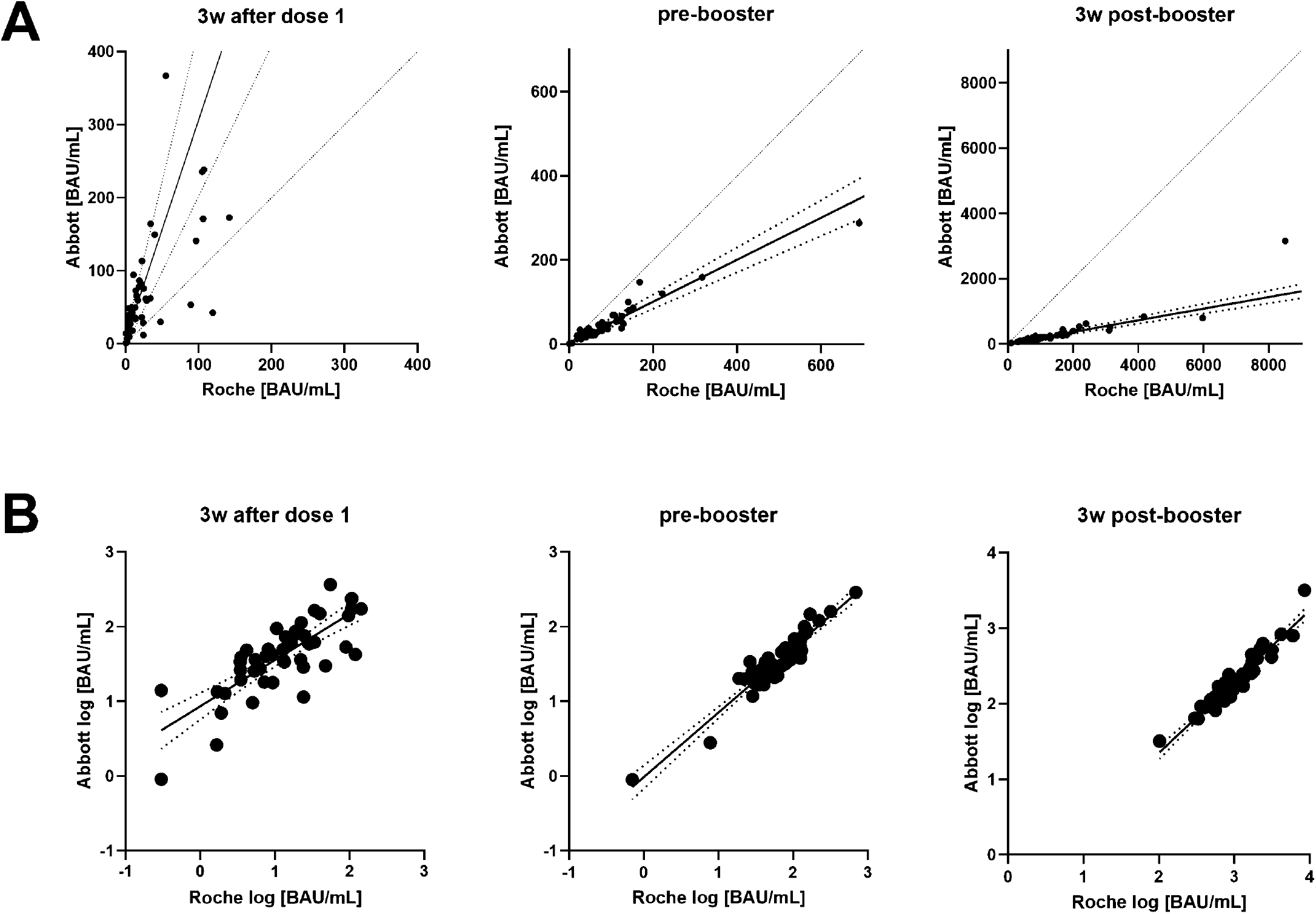
**A** Passing-Bablok regression for Roche and Abbott results; readings were converted to BAU/mL. The dotted lines are the 95% confidence intervals (CI) for the regression lines. The dashed lines represent lines of equality. **B** Linear regression (±95% CI) of logarithmic results from Roche and Abbott. 3w… 3 weeks; BAU/mL… binding antibody units per milliliter

### Correlation between binding assay and sVNT results

Next, we aimed to determine which of the two binding assays correlated better with neutralizing antibodies, particularly 3 weeks after the first dose, where the agreement between Roche and Abbott results was poorest. Neutralizing antibodies were estimated using the CE-IVD labeled cPass sVNT, with 30% inhibition as the threshold for positivity. 3w after the first dose, 6/50 (12%) participants yielded results below this threshold, with a median inhibition of 47.1 % (35.5-60.6). At week 11, directly before the booster, 4 of them rose above 30% inhibition, but two with initially positive results decreased below the threshold, resulting in a total of 4 individuals (8%) below 30% inhibition. The median neutralizing capacity remained nearly unchanged at an inhibition of 49.0 % (35.2-62.4). 3w after the booster dose, all but one participant presented with at least medium neutralizing capacity (>60%, see Fig. 2). The median increased to 95.2 % inhibition (92.1-97.9) at week 3 post booster; see Tbl. 1 and Fig. 2b.

As shown in Fig. 4, sVNT percent inhibition at 3 weeks after the first dose correlated with the Abbott assay at ρ=0.887, P<0.0001; in contrast, the correlation with the Roche test was slightly lower at ρ=0.666 (P<0.0001). At 11 weeks after the first dose, sVNT results correlated very well with both assays (Abbott: ρ=0.930, Roche: ρ=0.894, both P<0.0001). Similar results were observed at 3 weeks after the booster (Abbott: ρ=0.877, Roche: ρ=0.837, both P<0.0001).

**Fig. 4:**
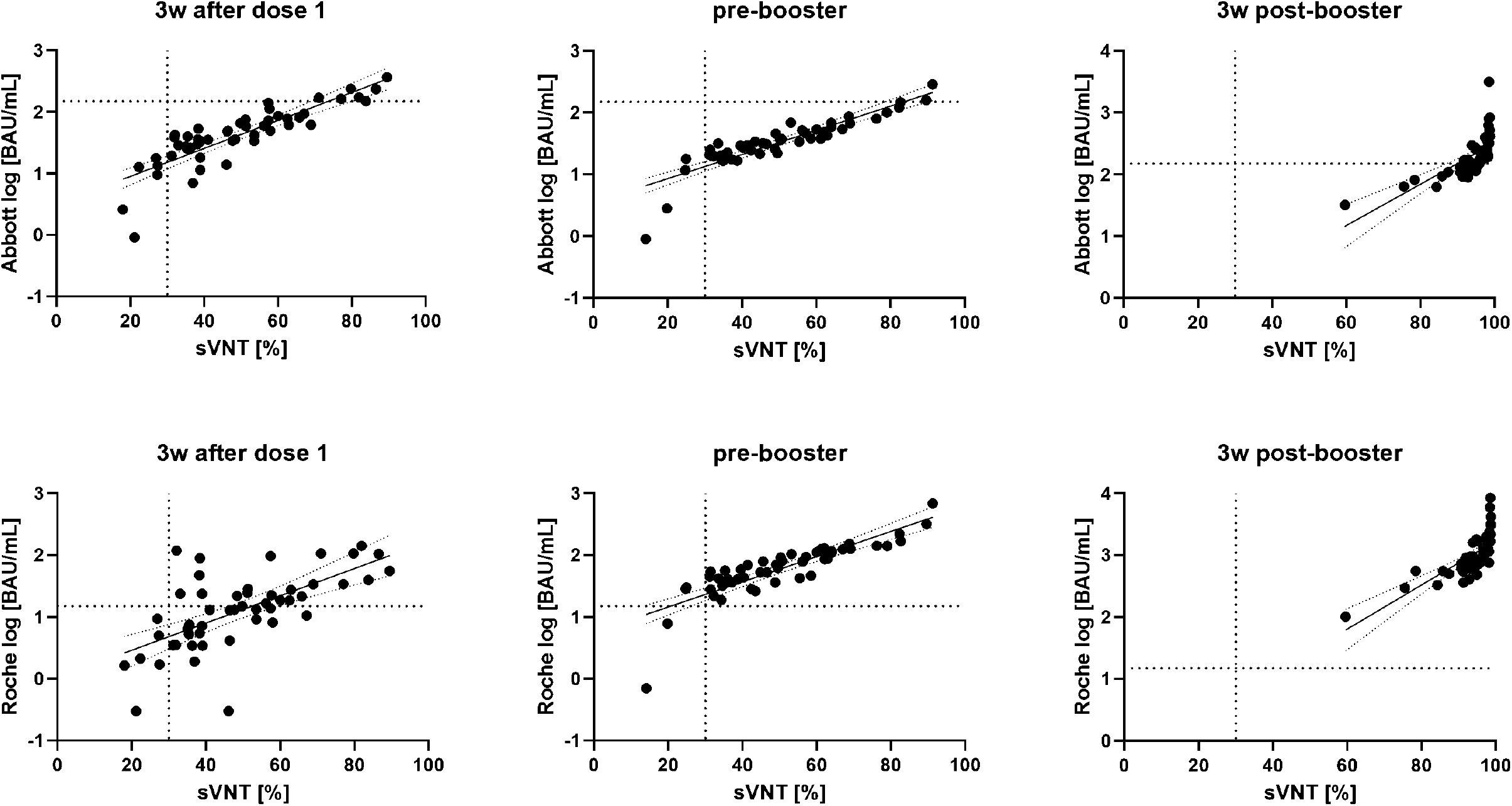
Linear regression lines (±95% confidence intervals) for c-pass surrogate virus neutralization test (sVNT) results and logarithmic binding assay results (top panel: Abbott, bottom panel: Roche). The dotted vertical line represents the sVNT’s threshold for positivity (30% inhibition). According to the manufacturers, Roche results ≥15 BAU/mL correlate with a positive neutralization test, Abbott results ≥149,1 BAU/mL correspond to a neutralization titer of at least 1:80 as indicated by horizontal dotted lines. 3w… 3 weeks; BAU/mL… binding antibody units per milliliter

These data suggest that qualitative differences between early and late SARS-CoV-2 antibodies may affect the comparability of serological tests.

### Relative changes of T-cellular responses, but not absolute IFNγ-levels, correlate with antibody levels

Finally, we examined the interactions between T-cellular and antibody responses (quantified with the Abbott and the Roche test). For this purpose, we compared the changes between the time before the booster (=11 after the first dose) and the time after the booster (3 weeks after the second dose). IFNγ-response to both used antigen mixtures (Ag1 and Ag2) increased after the booster shot: Ag1-Nil 0.10 IU/mL (0.05 – 0.16) to 0.15 IU/mL (0.09 – 0.32), P<0.0001; Ag2-Nil 0.16 IU/mL (0.09 – 0.31) to 0.31 IU/mL (0.16 – 0.98), see Fig. 3 and Tbl. 1. Levels from both antigen mixtures correlated well with each other (pre-booster ρ=0.725, P<0.0001; post-booster ρ=0.775, P<0.0001), see Fig. 5. Moreover, pre-booster levels were in good agreement with post-booster levels (Ag1-Nil ρ=0.786, P<0.0001; Ag2-Nil ρ=0.832, P<0.0001).

**Fig. 5:**
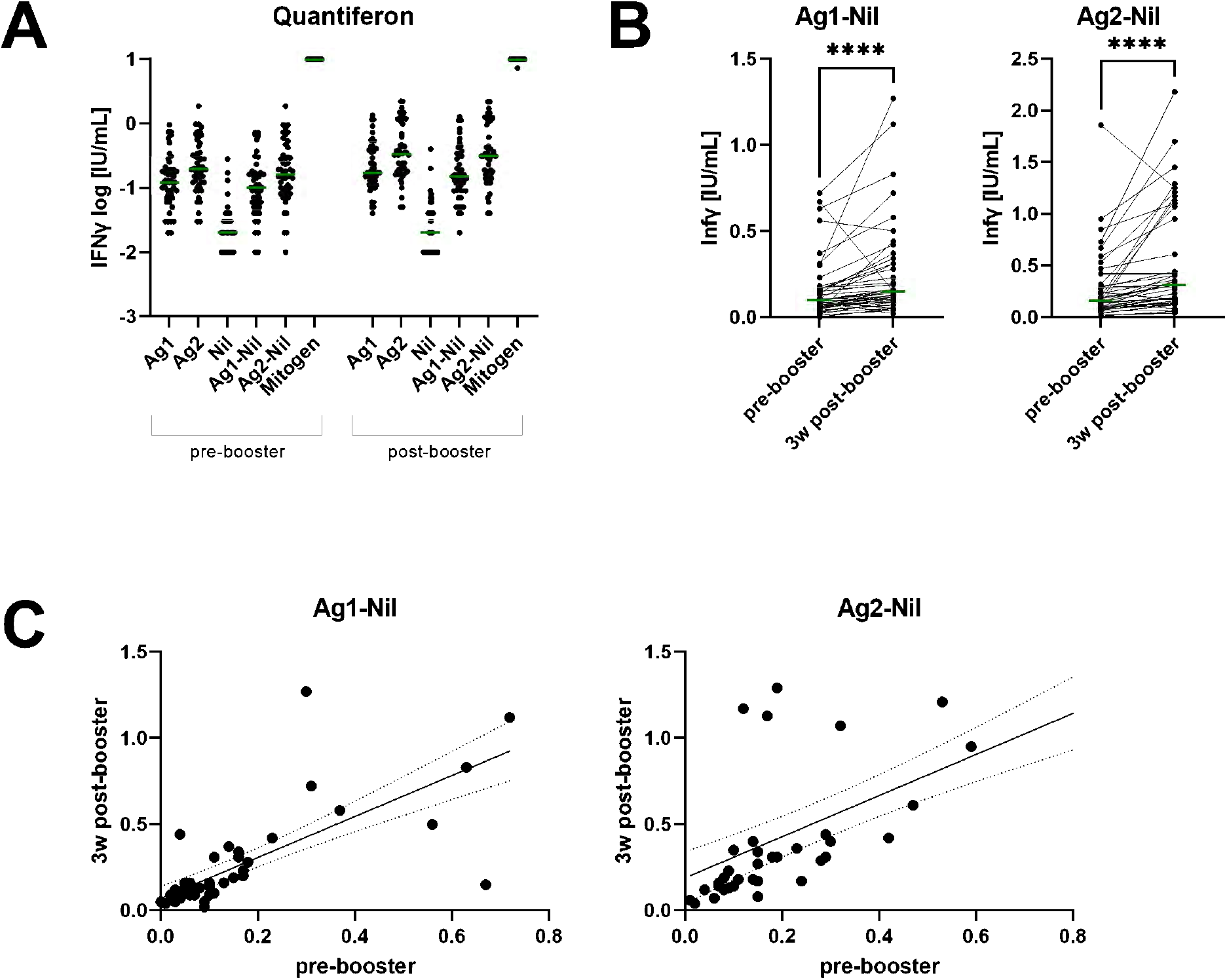
**A** Logarithmic IFNγ levels after stimulation of 1mL heparinized whole blood with Quantiferon SARS-CoV-2 antigen mixture 1 (Ag1), antigen mixture 2 (Ag2), Nil (negative control) and mitogen control, as well as Nil-corrected levels (Ag1-Nil, Ag2-Nil). Green lines indicate medians. **B** Pairwise comparisons of Ag1-Nil and Ag2-Nil in response to the booster shot. **** <0.0001. Green lines indicate medians. **C** Linear regression curves (±95% confidence intervals) of Ag1-Nil and Ag2-Nil before and after the booster shot. 3w… 3 weeks

Pre-booster interferon γ (IFNγ)-levels only weakly correlated with antibody levels. In fact, most correlations lacked statistical significance, and no relevant correlation was found 3 weeks after the booster. In contrast, the relative changes of cellular and binding assay antibody responses, calculated as 100*(post-booster – pre-booster)/pre-booster, correlated significantly after incubation with Ag2; however, for Ag1 statistical significance could not be reached (see Tbl. 2). Interestingly, no such correlation was observed for the sVNT.

**Tbl. 2:** Correlation Table. Spearman’s ρ of rank correlations between Nil-corrected interferon γ (IFNγ)-levels after incubation with Quantiferon SARS-CoV-2 antigen mixtures 1 (Ag1-Nil) or 2 (Ag2-Nil) and antibody levels. sVNT… surrogate virus neutralization test. % response… 100*(post-booster – pre-booster)/pre-booster

| **Pre-booster** | Abbott | Roche | sVNT |
| --- | --- | --- | --- |
| Ag1-Nil | 0.236 (*P*=0.100) | 0.258 (*P*=0.071) | 0.217 (*P*=0.130) |
| Ag2-Nil | **0.287 (*P*=0.044)** | 0.243 (*P*=0.089) | 0.252 (*P*=0.077) |
| **Post-booster** | Abbott | Roche | sVNT |
| Ag1-Nil | 0.037 (*P*=0.796) | 0.101 (*P*=0.485) | -0.001 (*P*=0.995) |
| Ag2-Nil | 0.067 (*P*=0.642) | 0.095 (*P*=0.510) | 0.024 (*P*=0.868) |
| **% response** | Abbott | Roche | sVNT |
| Ag1-Nil | 0.251 (*P*=0.082) | 0.247 (*P*=0.087) | 0.196 (*P*=0.178) |
| Ag2-Nil | **0.333 (*P*=0.018)** | **0.325 (*P*=0.021)** | 0.164 (*P*=0.255) |

## Discussion

SARS-CoV-2 specific anti-spike protein assays have been and are still widely used for serological studies^57^. In contrast to seroprevalence studies, where discriminating between positive and negative is usually sufficient, quantitative results are needed to adequately describe the response to SARS-CoV-2 vaccines and, ideally, to find a protective correlate^19,58^. However, there is a major obstacle on the way to such a protection correlate: the need for comparability of quantitative measurement results of different SARS-CoV-2 antibody tests^10,11^. In the present work, we compared two commercially available and broadly used CE-IVD marked SARS-CoV-2 antibody assays (Roche and Abbott). Both assays quantitate antibodies directed against the RBD domain of the SARS-CoV-2 spike protein and were referenced against the first WHO standard for SARS-CoV-2 antibodies, thus providing results in BAU/mL. We demonstrated in a previous study after vaccination with BNT162b that despite the standardization of SARS-CoV-2 antibody assays according to the first WHO standard for SARS-CoV-2 antibodies, the numerical values of different test systems are not interchangeable^27^. In the present work, we show for the first time that the problem of comparability is even more profound because the conversion of results may change dramatically with the time interval from vaccination.

Using samples from 50 individuals vaccinated with AZD1222, we could show that both assays detected SARS-CoV-2 specific antibodies in all but two participants 3 weeks after the first dose. Both non-responders were taking immunosuppressive drugs, which numerous studies have shown can lead to a decreased to absent response to the vaccine^59–61^. However, after the booster dose, the antibody levels markedly increased and reached detectable levels in all participants (Table 1 and Figure 2A). So, in contrast to people with previous COVID^62–65^, the second dose was required in our SARS-CoV-2 naïve population (negative for anti-nucleocapsid antibodies at all timepoints) to induce high antibody levels. The median relative change of individual antibody levels 3 weeks after the first versus 3 weeks after the second dose was nearly 20-times higher for Roche than for Abbott (80.8 vs. 4.5-fold change); see also Figure 2B. Despite targeting the same antigen (RBD) and converting to the same units (BAU/mL) using the first WHO standard for SARS-CoV-2 antibodies, not even the relative increases in antibody levels turned out to be comparable. The limited comparability of serological assays after vaccination is not specific for the AstraZeneca vaccine, as it was also observed after immunization with Pfizer/BioNTech BNT162b2^27^.

Furthermore, when looking at the difference between 3 and 11 weeks after the first vaccination dose, it was found that both the Abbott test and the cPass sVNT did not detect an increase in antibodies (Figure 2). In contrast, antibody levels measured by the Roche test increased >4-fold during this period. This discrepancy could be either explained by the inability of the Abbott and sVNT assays to distinguish such small changes in antibody concentration or the possibility that the Roche assay detects not only quantitative but also qualitative changes of the antibodies formed. Because previous studies failed to demonstrate a continuous increase in antibody levels for AZD1222 later than three weeks after vaccination, the Roche total antibody sandwich assay may also be sensitive to qualitative changes in nascent antibodies, in contrast to the Abbott IgG-specific assay^16,66^. This hypothesis is also supported by the observation that in direct comparison with the sVNT, the Roche assay underestimated inhibitory capacities at week 3 (Figure 4), which is discussed in detail below. Thus, overall, the assays studied show significant differences in the kinetics of antibody levels, which has been reported previously but was only rarely demonstrated from the same sample with different assays^28,67^. Although the correlation between Roche and Abbott improved over time, their relationship changed significantly depending on the time of blood sampling (Figure 3). At the first time point, Roche measured three times lower values in BAU/mL than the Abbott assay, 11 weeks after the first dose, Roche measured twice as high as Abbott, and finally, after the booster, Roche was median 5-6 times higher than Abbott.

As shown in numerous studies before, detection of SARS-CoV-2 anti-spike binding antibodies correlates well with the presence of functional neutralizing antibodies, so we wanted to examine differences between Roche and Abbott assays in this regard ^68–73^. The agreement between the results of the binding antibody assays and the neutralization test surrogate was generally good (Figure 4). In particular, at 11 weeks after the first vaccination, the correlation was excellent; after the booster, the correlation was technically limited due to many participants reaching the plateau of the sVNT. However, the worst correlation was found for the first antibody response 3 weeks after the first dose, and here Roche performed significantly worse than Abbott. This finding may be important because the improvement in Roche/sVNT correlation from ρ=0.666 to ρ=0.894 between 3 and 11 weeks after the first dose may indicate reduced sensitivity of the Roche assay for early antibodies. In other words, the discrepancy mentioned above that only Roche showed increasing antibody levels between 3 and 11 weeks after the first dose, while the other two tests showed identical or even slightly decreasing levels (Figure 2), could mean that the Roche test requires more matured antibodies to allow binding. It seems understandable that the binding of antibodies to two antigens, as necessary in the Roche sandwich test, places higher demands on the binding ability of antibodies than is the case in a typical anti-IgG-based detection method.

In line with previous studies, the second dose of AZD1222 substantially enhanced the initial antibody response in our cohort ^31,66,74^. Therefore, we wanted to investigate the relationship between the antibody levels and the cellular responses elicited by the booster. For this purpose, we used a SARS-COV-2 Quantiferon IFN-γ release assay similar to those known from tuberculosis diagnostics and compared the pre-booster and post-booster timepoints. As previously shown^75^, the second vaccination dose induced an increase in cellular reactivity (Figure 5). However, we found only weak, mostly statistically non-significant correlations between antibody levels and IFN-γ levels before the booster and no correlation at all after the booster (Table 2).

In contrast, the percent cellular response (fold change) to the booster correlated significantly with the percent antibody response (ρ=0.33 for both binding assays), see Table 2. This finding suggests that the increase of antibodies after a booster shot, which is detected by both binding assays, can be substantiated by an accompanying cellular reaction. In contrast, we found no correlation between the relative changes in cellular and sVNT response, which might be partly explained by the limited measurement range of the sVNT. However, since not all antibodies formed are functionally active neutralizing antibodies (NAbs), even not all of those specifically directed against the RBD domain of the spike protein, the binding antibodies may be superior to the measurement of NAbs here as a correlate for cellular activation.

This study has several strengths and limitations: although 50 participants might be considered a relatively small cohort, we have shown in previous work that this number is sufficient for such comparative approaches and that our data could be replicated in much larger cohorts^27,76^. One strength of our study is that we followed exact time points for blood sampling in the context of a prospective observational study. Furthermore, our cohort using AZD1222 (inducing significantly lower median antibody levels than, e.g., BNT162b2) has the advantage of a broader distribution of values across the measurable spectrum with a very low proportion of results above 1,000 BAU/mL. As this value represents the upper limit of referencing with the WHO SARS-CoV-2 antibody standard, a linear relationship is no longer guaranteed for values above this, leading to unwanted biases in comparing different antibody tests.

In summary, with the present work, we show for the first time that the comparability of quantitative anti-spike SARS-CoV-2 antibody tests is highly dependent on the timing of blood collection. Therefore, it does not seem feasible to compare different quantitative SARS-COV-2 antibody results without standardization of the time of collection.

## Data Availability

Data is available for interested researchers in compliance with the GDPR upon request from the corresponding author.

## Acknowledgments

We thank all sample donors for their valuable contributions. We further thank Martina Trella, Susanne Keim, Borka Radovanovic-Petrova, Monika Martiny, Jadwiga Konarski, Bernhard Haunold, Maedeh Iravany, and Shohreh Lashgari for perfect technical and administrative assistance. The MedUni Wien Biobank is part of the Austrian biobanking consortium BBMRI.at.

The Department of Laboratory Medicine received compensation for advertisement on scientific symposia from Roche and Abbott and holds a grant for evaluating an in vitro diagnostic device from Roche.

There was no specific funding received for the present work.

## Notes

### Funding Statement

No external funding was received.

### Author Declarations

The study protocol was reviewed and approved by the Medical University of Vienna ethics committee (1066/2021) and conforms with the Declaration of Helsinki.

